# Argon Laser Photocoagulation for Treatment of Presumed Trematode-induced Granulomatous Anterior Uveitis in Children

**DOI:** 10.1101/2020.12.30.20248850

**Authors:** AlahmadyHamad Alsmman, Abdelsalam Abdalla, Mohamed Ezzeldawla, Shaimaa Abd Elmatin, Mortada Ahmed Abozaid

## Abstract

**Background/Aims:** To assess the safety and efficacy of argon laser photocoagulation as a new modality for the treatment of presumed trematode-induced granulomatous anterior uveitis (PTGAU) in children.

**Methods:** Forty-eight eyes of 48 children with PTGAU with pearl-like nodule(s) in the anterior chamber were included in this prospective non-randomised controlled clinical trial. Patients were divided into two groups: group A (23 eyes) was treated with one session of argon laser applied to the anterior chamber nodules, and group B (25 eyes) received medical treatment in the form of topical steroid and cycloplegic eye drops with trans-septal triamcinolone injection.All cases were followed up for 3 monthswith measurement of VA, assessment of anterior chamber reaction, and size of the pearl-like nodules.

**Results:** In group A, 22 eyes (95.65%) showed regression of the pearl-like nodules with resolution of the anterior chamber reaction (flare and cells) and improvement in visual acuity from 0.52±0.12 to 0.06 ± 0.08logMAR (*p* value **<0**.**001)**. Such improvement was maintained within the 3-month follow-up period. In group B, 23 eyes (92%) showed initial regression of the granulomas,whichwas maintained in only 14 eyes (56%),with 9 eyes experiencing recurrence after 3 months of follow-up.

**Conclusion:** Argon laser photocoagulation is a safe and effective novel treatment for PTGAU with pearl-like nodules in the anterior chamber in children. Larger studies with longer follow-up periods are needed to confirm these results.

**PRECIS:** Argon laser photocoagulation is a novel non-invasive line of treatment that can be added to the armamentarium for presumed trematode-induced granulomatous anterior uveitis in children.

## INTRODUCTION

Presumed trematode-induced granulomatous anterior uveitis (PTGAU) is a relatively common anterior uveitis among paediatric patients from rural Egypt. It has been linked to swimming or bathing in the Nile river and freshwater canals.[1,2]It is characterised by chronic granulomatous iridocyclitis with one or more pearl-like nodules (pearl tumour) of variable sizes and locations within the anterior chamber (AC) with or without subconjunctival nodules. Most cases occur in menwho present with acute exacerbations of iridocyclitis, resulting in complications thatthreatenvision, such as synechiae, cataract, glaucoma, and/or phthisis bulbi. In addition, untreated granulomas usually heal by fibrosis, leaving a permanent scar, which can be visually significant.[3-6]Molecular evidence of digenic trematode as a cause of this distinct form of uveitis was reported in a recent study.[7]

Schistosomiasis or bilharziasis is an endemic disease in Egypt. It is caused by the *Schistosoma* genus (flat trematodes or blood flukes), which can induce granulomatous reactions in ocular tissues around the eggs or adult worms. *Schistosoma haematobium* is the predominant species in Upper Egypt, while *S. mansoni*, which is predominant in lower Egypt, was isolated from the AC in several cases reported.[8-11]

The treatment options for this granulomatous uveitis include topical steroids and cycloplegics, trans-septal steroids, systemic steroids and anti- parasitics, limbal cryotherapy,[12] and surgical excision.[1] However, many children may develop complications from both conservative treatments, especially steroids [13]or surgical intervention.

Since its invention in 1964 by William Bridges, the argon laser has been used in a number of ophthalmic conditions becauseof its precision in targeting the intended structures, especially in glaucoma and retinal photocoagulation procedures. In addition, it is used in many dermatologic lesions such as ulcers and polyps.[14]

In this non-randomised controlled clinical trial, we tried a new modality of treatment in the form of argon laser photocoagulation of the pearl-like nodules in the AC and compared the results to a control group of medical treatment. In the absence of animal models, we relied on a pilot set of data formed fromthree cases to confirm the safety and efficacy of the procedure before proceeding with the clinical trial.

## PATIENTS &METHODS

This study was conductedbetweenMay andOctober 2020 at the ophthalmology department of Sohag University Hospital in collaboration with Nour-Eloyoun private centre in Sohag, Egypt. The study followed the tenets of the Declaration of Helsinki and was approved by the ethical committee of the Sohag Faculty of Medicine. The study was also registered as a clinical trial in the Pan African Clinical Trial Registry with the number PACTR202008516974176. After explanation of the procedure’s benefits and risks, written informed consent was obtained from the parents of the children.

Forty-eight children (< 18 years) with active PTGAU were included in the study. The inclusion criteria were as follows: one or two pearl-like masses in the AC, flare and cells ≥ +2 according to the Standardised Uveitis Nomenclature (SUN) classification, anda visual acuity better than 6/60 in the affected eye.

The exclusion criteria of this study included presence of inactive scarred membranous lesions, previous diagnosis of cataract, glaucoma, or phthisis bulbi, and a history of ocular trauma or surgery. Patients with a known cause of uveitis or systemic granulomatous disease were also excluded.

All patientsunderwent a complete ophthalmological evaluation including detailed history about the onset and course of the iritis and its relation to river water exposure. Ocular examination was performed via slit lamp evaluationand included documentation ofthe number and size of pearl-like lesions as well as grading the ACreactionofcells and flareaccording to the SUN nomenclature.[15]Visual acuity (corrected and uncorrected) and intraocular pressure(IOP) were measured, and a fundus examination was also completed. A systemic work-up was performed in all cases in the form of stool and urine analysis, chest X-ray, complete blood count (CBC), erythrocyte sedimentation rate (ESR), and anti-bilharzial antibody titre.

The children were divided into two groups according to the consent of their parents; those who agreed to surgical intervention in the form of one session of argon laser were included in group A (23 eyes), while those who refused such treatment were included in group B (25 eyes).

### Surgical technique in group A

Under topical anaesthesia, all cases received one session of argon laser photocoagulation applied directly to the pearl-like lesions in the AC through the argon laser lens. The settings of the argon laser included:100 to 200 µm spot size;0.2 to 0.4 seconds duration;250 to 400 mJ power; and 10 to 25 shots, depending on the response of the lesion (blanching and shrinkage) as shown in Figure1. Cases were performed by two surgeons (AA and MA).

**Figure1.**
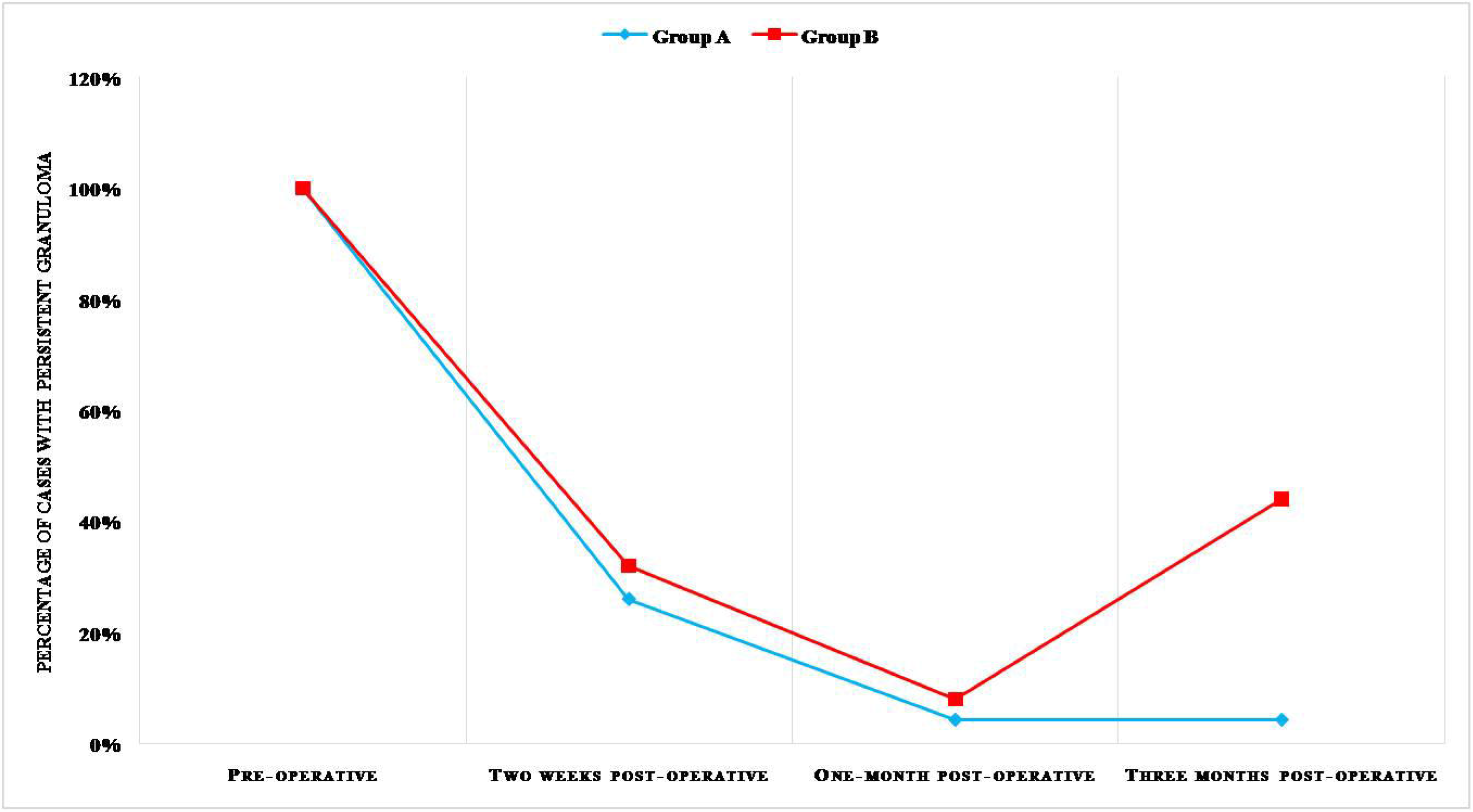
A case from group A with single Ac granuloma at 6 O’clock position.(a) Before,(b) immediately after,and (c) three-months after argon laser photocoagulation.

### Perioperative treatment

Topical steroids and cycloplegics were started 3 days before the session. The following topical eye drops were used postoperatively: gatifloxacin 0.3% five times daily for oneweek,prednisolone acetate 1% five times daily with gradual taper over4 weeks, cyclopentolate hydrochloride three times daily for 3 days, and brimonidine twicedaily for one week.

### Medical treatment in group B

Patients whose parents refused the surgical intervention were given the following conservative regimen:

1. Trans-septal (orbital floor) triamcinolone acetonide (40 mg/mL) injection under topical anaesthesia after reassurance of the patients.
2. Topical prednisolone acetate 1% eye drops:five times daily with a gradual taper over 4 weeks.
3. Cyclopentolate hydrochloride 1%: three times daily for 2 weeks.
4. Topical gatifloxacin 0.3% eye drops:five times daily for one week (prophylactic use to prevent secondary bacterial conjunctivitis).

### Statistical analysis

Data wereanalysed using IBM SPSS Statistics for Windows version 20.0 Quantitative data were expressed as median and inter-quartileranges. Qualitative data were expressed as numbers and percentages. The data were tested for normality using the Shapiro-Wilk test. An independentsamples test was used for normally distributed data. The nonparametric Mann–Whitney test and Friedman test were used for data thatwere not normally distributed.

Chi-square (χ ^2^) and Fisher’s exacttests were used for comparison of qualitative variables as appropriate. The Friedman test was used for comparison between repeated AC cell measures of the studied patients. Cochran’s Qtest was used for comparison between repeated granuloma examinations of the studied patients. A 5% level was chosen as the level of significance in all statistical tests used in the study.

## RESULTS

This clinical trial included 48 eyes of 48 children with PTGAU and pearl-like nodules in the AC. Cases were divided into two groups: group A included 23 eyes and was treated by one session of argon laser applied to the AC nodules, and group B included 25 cases and received conservative medical treatment. The two groups showed no differenceswith regard to their baseline demographics or ocular findings, as shown in Table1.

**Table -1.**
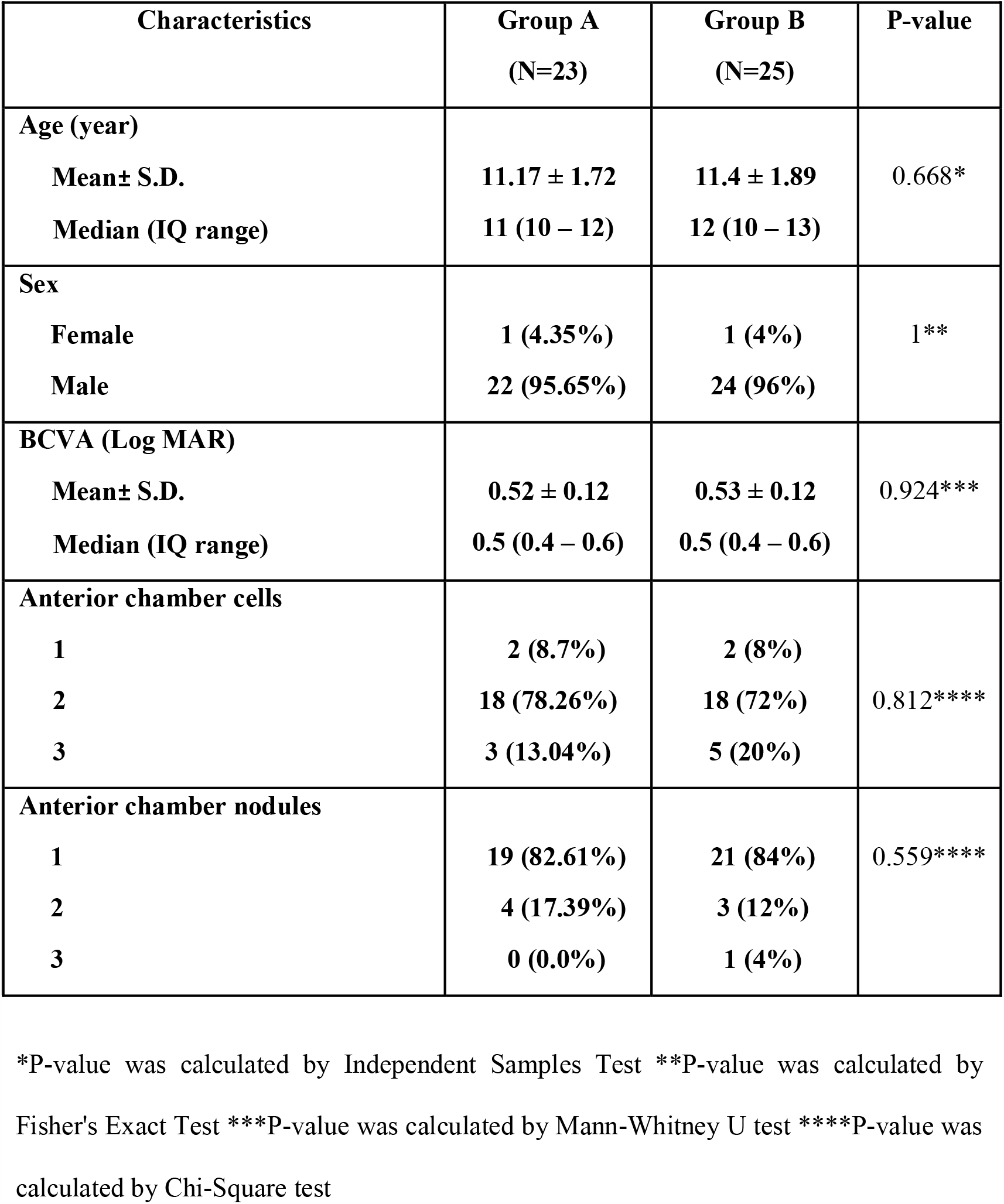
comparison between the two study groups regarding their preoperative measures.

The results of systemic investigations were irrelevant, with antibilharzial antibodies being positive in 12 cases from group A and 10 cases from group B.The mean age of group Awas 11.17 ± 1.72 years, ranging from 8to14 years, while that of group B was 11.4 ± 1.89 years,ranging from 8 to 14 years. Most children were boyswith a history of swimming or bathing in the Nile river,which can be explained by the cultural aspects thatprevent girls from bathing or swimming in the water of the Nile riveror its canals.

The preoperative mean BCVA in group A was 0.52±0.12logMAR, while the postoperative mean BCVA was 0.22 ± 0.1, 0.11 ± 0.08, and 0.06 ± 0.08 after 2 weeks, one month, and 3 months, respectively (p value < 0.001). The number of cases with +2 and +3 AC cells decreased from 21 preoperatively to 0 after 3 months of follow-up (Table2). The AC nodule(s) shrunk in22 cases (95.65%), with resolution of the AC reaction within 2-4 weeks.This response was maintained through the 3-month follow-up period (Table3).

**Table-2.**
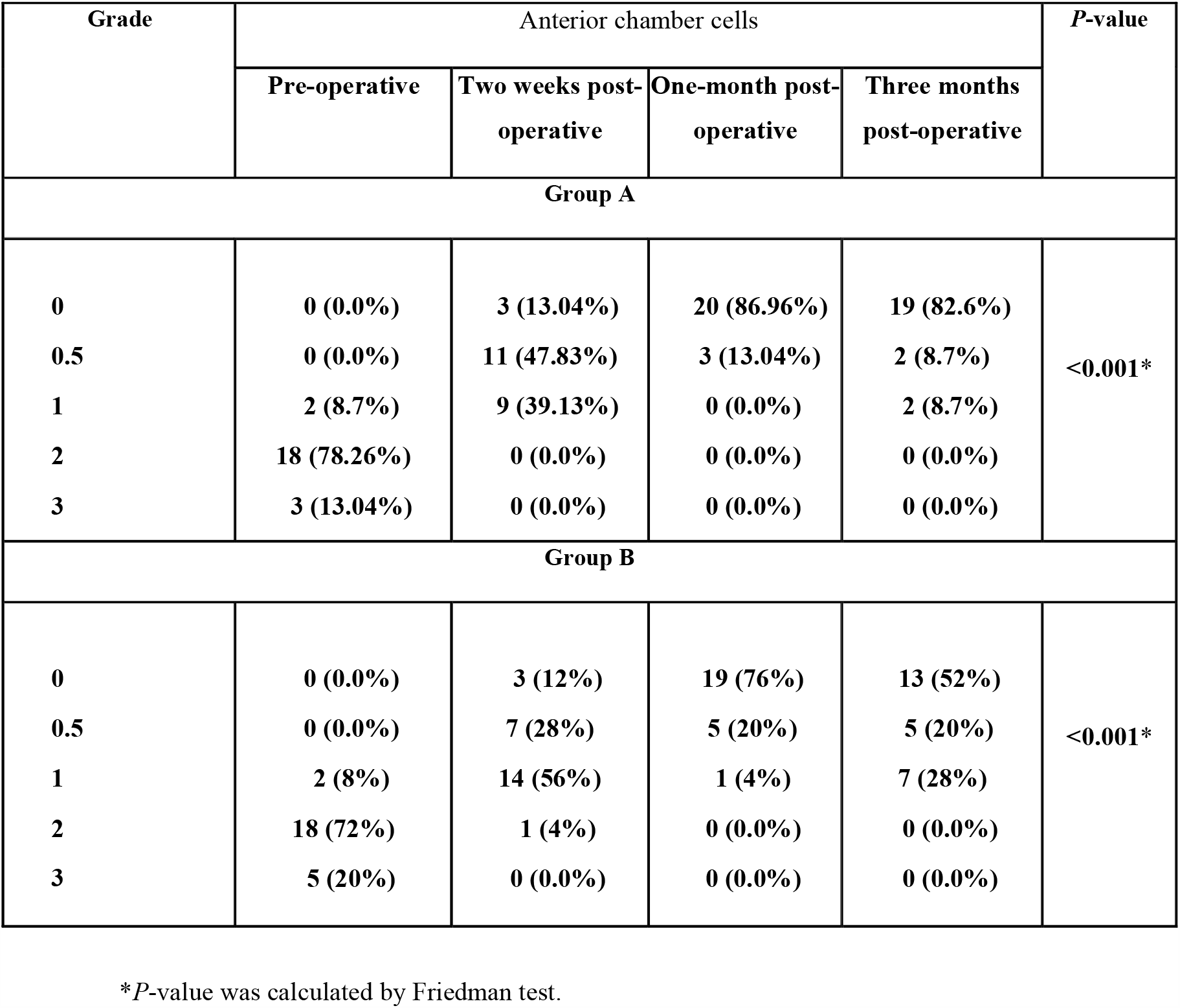
Comparison between the repeated examinations of AC cells in the 2 groups.

| Grade | Anterior chamber cells |  |  |  | P-value |
| --- | --- | --- | --- | --- | --- |
|  | Pre-operative | Two weeks post-operative | One-month post-operative | Three months post-operative |  |
| Group A |  |  |  |  |  |
| 0 | 0 (0.0%) | 3 (13.04%) | 20 (86.96%) | 19 (82.6%) | <0.001* |
| 0.5 | 0 (0.0%) | 11 (47.83%) | 3 (13.04%) | 2 (8.7%) |  |
| 1 | 2 (8.7%) | 9 (39.13%) | 0 (0.0%) | 2 (8.7%) |  |
| 2 | 18 (78.26%) | 0 (0.0%) | 0 (0.0%) | 0 (0.0%) |  |
| 3 | 3 (13.04%) | 0 (0.0%) | 0 (0.0%) | 0 (0.0%) |  |
| Group B |  |  |  |  |  |
| 0 | 0 (0.0%) | 3 (12%) | 19 (76%) | 13 (52%) | <0.001* |
| 0.5 | 0 (0.0%) | 7 (28%) | 5 (20%) | 5 (20%) |  |
| 1 | 2 (8%) | 14 (56%) | 1 (4%) | 7 (28%) |  |
| 2 | 18 (72%) | 1 (4%) | 0 (0.0%) | 0 (0.0%) |  |
| 3 | 5 (20%) | 0 (0.0%) | 0 (0.0%) | 0 (0.0%) |  |

\**P*-value was calculated by Friedman test.

**Table-3.** Comparison between the repeated Granuloma examinations in the two groups.

| Characteristics | Granuloma |  |  | P-value |
| --- | --- | --- | --- | --- |
|  | Two weeks post-operative | One-month post-operative | Three months post-operative |  |
| Group A |  |  |  |  |
| Resolved | 17 (73.91%) | 22 (95.65%) | 22 (95.65%) | 0.007* |
| Unresolved | 6 (26.09%) | 1 (4.35%) | 1 (4.35%) |  |
| Group B |  |  |  |  |
| Resolved | 17 (68%) | 23 (92%) | 14 (56%) | 0.001* |
| Unresolved | 8 (32%) | 2 (8%) | 11 (44%) |  |

\*P-value was calculated by Cochran's Q test

No serious complications were detected during or after the argon laser session. Only one case showed an increasein IOP to 27 mmHg the day after the argon laser session;this casewas successfully managed with a topical beta-blocker. In addition, hyphaema was noticed in one case immediately after the session (figure 2) and resolved completely within 5 days. Only one case in group A showed resistance to photocoagulation with minimal initial shrinkage followed by recurrence within 3 months. This case is now under medical treatment untilthe arrangement of a second session of the argon laser.

**Figure2.**
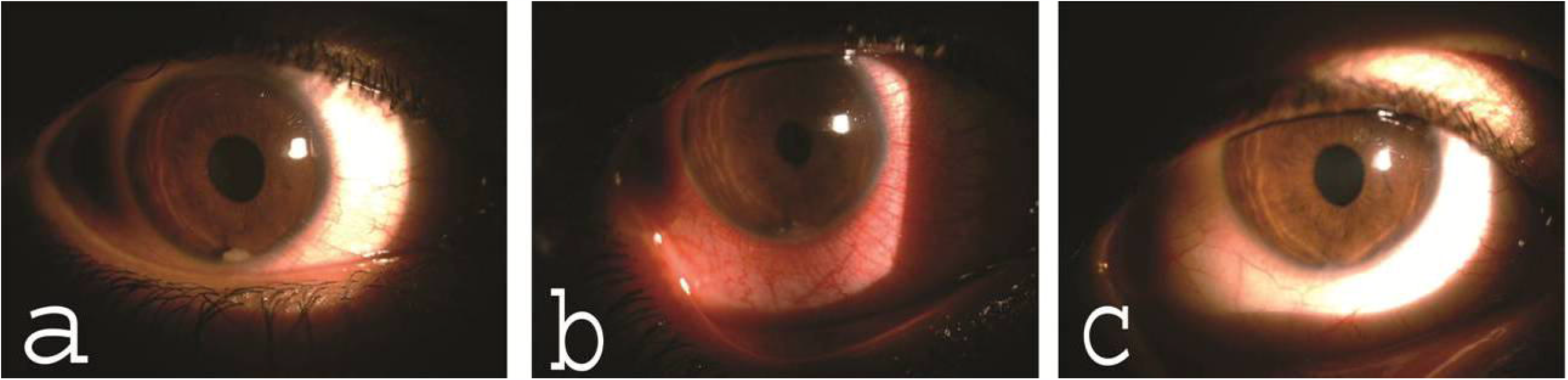
A case from group A with single AC granuloma at the 3 O’clock position that was complicated by postoperative hyphaema.(a) Before and (b) immediately after argon laser photocoagulation.

In group B the preoperative mean BCVAwas 0.53±12(logMAR), while the postoperative mean BCVA was 0.24 ± 0.01, 0.1 ± 0.07, and 0.16 ± 0.17 after 2 weeks, one month, and 3 months, respectively (p value **<0**.**001)**. The number of cases with +2 and +3 AC cells decreased from 23 preoperatively to 0 after 3 months of follow-up; however, cases with +0.5 and +1 increased from 2 to 12 in the same time period(Table2). The AC nodule(s) shrunk in 23 cases (92%) within one month after medical treatment with resolution of uveitis. However, 9of these 23 casesshowed recurrence by the end of 3 months, with reappearance of the AC reaction and the percentage of nodules that had disappeared decreasing to only 56%,as seen in Table3 and Figure 3.

**Figure3.**
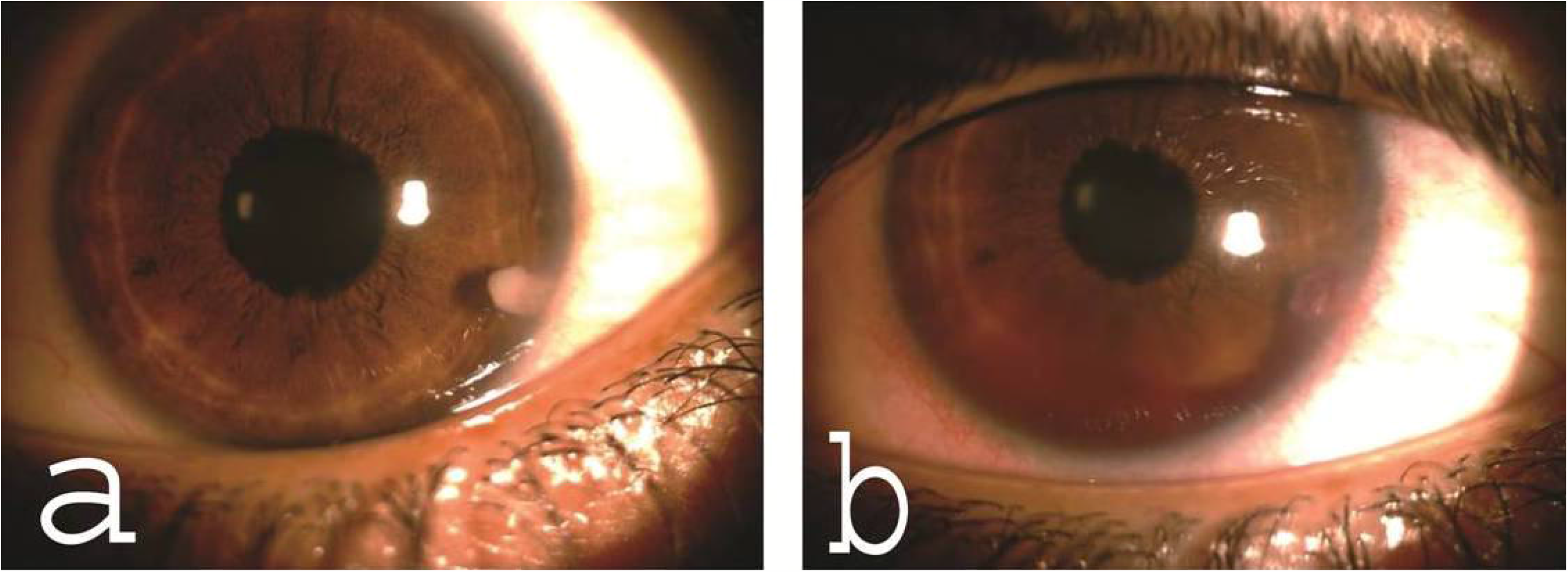
A rise in the percentage of persistent granulomas after initial decline in group B compared to a maintained decline demonstrated in group A.

No complications were reported from the trans-septal triamcinolone injection, and no cases of cataract or steroid induced glaucoma were detected.

## DISCUSSION

Schistosomiasis or bilharziasis has been endemic in Egypt for thousands of years andis associated with many life-threatening complications, such as hepatic failure and cancer of the urinary bladder. Thanks to the antiparasitic praziquantel and mass education about the seriousness and transmission of the disease, bilharziasis has been brought under control over the last few decades. However, Upper Egypt (the southern part of Egypt) still suffers from a relatively high infestation rate, mostly due to its agricultural environment and the low socio-economic status of the inhabitants.

PTGAU is a potentially blinding ocular condition associated with bathing or swimming in local river water, such as that of the Nile river in Egypt. *Schistosoma haematobium* is the most likely cause of this condition in Upper Egypt.

Many children with this disease do not respond well to topical or systemic medical treatment andthus require more aggressive forms of treatment, such as surgical excision, with potential complications including those of general anaesthesia.

In this pilot study, we reported the safety and efficacy of argon laser photocoagulation of the pearl-like nodules in the AC of cases with PTIGAU. Forty-eight eyes of 48 children with PTIGAU associated with one or two pearl-like nodules in the AC were included in this clinical trial. Cases were divided into two groups: the first group or group Awas treated with one session of argon laser photocoagulation applied to the pearl-like nodule(s) with peri-operative topical steroids and cycloplegics. The second group or group B was treated conservatively with trans-septal steroids, topical steroids, and cycloplegics. After 3 months of observation, a statistically significant difference in the outcome was detected between the two groups, with marked improvement in the argon laser groupas detected bybetter VA, better control of inflammation, and lower rate of recurrence (p value 0.002).

The possible mechanism of action of the argon laser is the destruction or thermal ablation of the pearl-like nodules in the AC,which are associated with the trematode larvae or adult worm, thus eliminating the antigenic stimulant of the relapsing granulomatous anterior uveitis.

In their prospective case series, El Nokrashy et al. [16] assessed the efficacy of both systemic antiparasitic treatment alone and that of the systemic antiparasitic treatment combined with surgical aspiration in 30 eyes of 30 children with PTGAU presenting with AC granuloma. They concluded that antiparasitic treatment alone (praziquantel+metronidazole) is effective for small granulomas, while surgical aspiration is a good adjuvant treatment for large granulomas.

Amin et al. [17]reported the clinical prescription and laboratory analysisof 110 children with PTGAU in Egyptian children. Surgical excision of the lesions was performed in 14 cases. Histopathological examination of these lesions after removal revealed granulomatous inflammation formed by central suppuration surrounded by epithelioid cells and sheets of lymphocytes. Polymerase chain reaction analysis performed for six of the 14 samples showed positive results for trematode DNA.

Sadek et al. [18]compared the outcomes of medical treatment to surgical intervention (excision of granuloma and AC wash) in 41 eyes of 39 patients with active infection, defined as flare and cells ≥ +2 and the presence of a white AC granuloma > 3 mm.

After two weeks, the granuloma disappeared in 20 of the 21 eyes in the surgical groupversus only two eyes of the 20 eyes in the conservative treatment group. They concluded that surgical excision of large granulomas results in a more rapid and complete cure.

To the best of our knowledge, this study is the first to test the use of argon laser photocoagulation in the treatment of PTIGAU. The technique is simple, cheap, non-invasive, and widely available in the developing countries.While the results are encouraging, future studies with larger samples and longer durations of follow-up are needed to confirm the results.

## Data Availability

data will be available in a repository immediately after publication

## Acknowledgements

We would like to thank Editage (www.editage.com) for English language editing.

## Competing Interests

The authors do not have any conflict of interest regarding the procedures or medications used in this study.

## Funding

**None**

## Data availability statement

All data relevant to the study are uploaded as supplementary information.

## Notes

### Competing Interest Statement

The authors have declared no competing interest.

### Clinical Trial

PACTR202008516974176
https://pactr.samrc.ac.za/Researcher/ManageTrials.aspx

### Author Declarations

Ethics committee of Sohag faculty of medicine IBR#S20-131

